# Bilingualism’s protective effects in Alzheimer’s disease: Mechanisms of resilience and resistance

**DOI:** 10.64898/2026.02.13.26345903

**Authors:** Wenfu Bao, Stephanie M. Grasso, Isabel Sala, M. Belén Sánchez-Saudinós, Judit Selma-González, Javier Arranz, Nuole Zhu, Sara Rubio-Guerra, Íñigo Rodríguez-Baz, María Carmona-Iragui, Isabel Barroeta, Ignacio Illán-Gala, Juan Fortea, Olivia Belbin, Lídia Vaqué-Alcázar, Marco Calabria, Eider M. Arenaza-Urquijo, Alexandre Bejanin, Daniel Alcolea, Alberto Lleó, Miguel Ángel Santos Santos

**Affiliations:** Department of Speech, Language, and Hearing Sciences, The University of Texas at Austin, Austin, Texas, USA; Sant Pau Memory Unit, IR SANT PAU, Hospital de la Santa Creu i Sant Pau, Barcelona, Spain; Centro de Investigación Biomédica en Red en Enfermedades Neurodegenerativas (CIBERNED), Madrid, Spain; Institut de Neurociències, Universitat Autònoma de Barcelona, Barcelona, Spain; Barcelona Down Medical Center, Fundació Catalana Síndrome de Down, Barcelona, Spain; Department of Medicine, Faculty of Medicine and Health Sciences, Institute of Neurosciences, University of Barcelona, Institut d’Investigacions Biomèdiques August Pi i Sunyer (IDIBAPS), Barcelona, Spain; NeuroADaS Lab, Universitat Oberta de Catalunya, Barcelona, Spain; Barcelona Institute for Global Health (ISGlobal), Barcelona, Spain

**Author notes:** **Corresponding author**: Miguel Ángel Santos Santos. Hospital de la Santa Creu i Sant Pau, Sant Antoni Maria Claret, 167, 08025 Barcelona, Spain.

**Keywords:** Alzheimer’s disease, bilingualism, biomarker, cognition, resilience, resistance

## Abstract

**INTRODUCTION:** Bilingualism is among several lifestyle factors associated with protection against cognitive decline, yet the biological mechanisms through which it exerts these effects remain poorly understood.

**METHODS:** We compared neuropsychological functioning and biofluid markers of brain health between active (*n* = 280) and passive (*n* = 287) Spanish-Catalan bilinguals with biomarker-confirmed Alzheimer’s disease (AD).

**RESULTS:** Active bilinguals outperformed passive bilinguals on tests assessing attention/executive functions, language, and visuospatial/visuomotor functioning, demonstrating resilience given the same AD biological stage across participants. Active bilinguals also exhibited significant differences in cerebrospinal fluid and plasma biomarkers of amyloid burden and neuroinflammation, suggesting both resilience and resistance to AD pathophysiologic mechanisms.

**DISCUSSION:** The protective effects of bilingual experience may engage both resilience and resistance to AD pathophysiology mechanisms. These results underscore the importance of capturing bilingualism in aging cohorts and the study of how lifestyle and sociocultural factors shape the biological expression of neurodegenerative disease.

## 1. Background

The protective effects of bilingualism have been observed across typical and pathological aging [1–3]. Despite mixed findings, many studies report improved cognitive performance and greater neurostructural integrity in bilinguals compared to monolinguals [1,4,5]. Evidence also links bilingualism to a delayed dementia onset of up to five years [2,6]. However, prior research has focused on participants without biomarker confirmation; clinical diagnoses of Alzheimer’s disease (AD) or mild cognitive impairment (MCI) based solely on symptoms and cognitive tests are often inaccurate [7]. Moreover, relative to other cognitive reserve and/or resilience factors (e.g., education, sex [8–11]), the biological mechanisms underlying bilingualism’s protection have received less attention. Examining fluid biomarkers may reveal whether bilingualism influences cognitive decline by conferring resilience or resistance to AD pathology.

Nevertheless, research on the biological basis of bilingualism’s effects against cognitive decline remains nearly nonexistent except for two adjacent studies [12,13]. Notably, little work has studied individuals at the clinical dementia stage. Estanga et al [12] assessed cerebrospinal fluid (CSF) AD biomarkers in cognitively unimpaired participants and found lower CSF total tau (t-tau) in early bilinguals compared to monolinguals. This provides initial evidence that early bilingualism is associated with a more favorable CSF AD biomarker profile, suggesting resistance to tau pathology. In contrast, Grasso et al [13] observed a disadvantageous plasma biomarker profile in bilinguals with MCI who were impaired in verbal fluency, indicated by lower amyloid-beta (Aβ) 42/40 levels relative to bilinguals with MCI but unimpaired performance on verbal fluency.

This study investigates the effects of bilingualism on neuropsychological functioning and biofluid (i.e., CSF and plasma) markers of brain health, spanning AD-specific pathophysiologic processes (Aβ and tau accumulation) and non-specific neurodegeneration (e.g., neurofilament light or NfL) and neuroinflammation (e.g., glial fibrillary acidic protein, GFAP). Leveraging a cohort [14] with CSF biomarker-confirmed AD (Aβ and tau positive, A+T+ [15]) categorized into active vs. passive bilinguals (further described below), we explore the biological mechanisms, through the lens of resilience and resistance, that may subserve the bilingualism’s impact.

In the context of AD, consistent with previous frameworks [16–18], we operationalize resilience as better-than-expected cognition in relation to pathology or higher-than-expected pathology given cognition, and resistance as absent or lower-than-expected pathology given age, genetics, disease stage, etc. We note that definitions surrounding reserve, resilience, and resistance have evolved and investigators may subscribe to different models (e.g., [19,20]). Yet, our aim is not to adjudicate between these frameworks, but to apply the aforementioned definitions of resilience and resistance in interpreting bilingualism’s effects on AD.

Both education and biological sex are related to dementia risk [8]. Higher education has been identified as a resilience factor, given its association with increased Aβ deposition via positron emission tomography (PET) in AD compared to low-educated participants matched for cognitive severity [9]. Regarding sex differences, women have higher dementia incidence rates [8] and generally perform better on verbal tests, whereas men tend to outperform on visuospatial tasks [10] and show elevated CSF NfL levels [11]. Evidence further suggests that bilingualism may modulate sex differences in cognition and biomarkers [21–23].

Therefore, we hypothesize that bilingualism might interact with sex or education to confer resilience and resistance, reflected in neuropsychological and biomarker outcomes. As participants were at the same AD biological stage (A+T+), we consider better neuropsychological performance and/or greater pathology (measured via biofluid levels) as evidence for resilience, and less pathology as evidence for resistance. Given the influence of lifestyle factors on AD pathology [8,9], we also hypothesize that bilingualism-related cognitive differences may be explained by biological variability, and thus explore the mediating role of biomarker levels to probe potential biological pathways underlying the bilingualism-cognition link.

## 2. Methods

### 2.1. Participants

We included 567 Spanish-Catalan bilinguals (346 female, mean age = 75.8 ± 4.59 years) from the Sant Pau Initiative on Neurodegeneration (SPIN) cohort [14] at Sant Pau Memory Unit in Barcelona. All participants had a syndromic diagnosis of MCI or AD dementia (see [14] for clinical criteria), with the age at symptom onset ≥ 65 years. Additionally, their CSF biomarkers supported AD pathophysiology (A+T+; see [24] for detailed cutoffs). Participants were classified as active bilinguals if they reported actively using both Spanish and Catalan at the clinical visit, and they were tested in their preferred language (Spanish or Catalan) during neuropsychological evaluation. Individuals reporting active use of Spanish only and passive exposure to Catalan were classified as passive bilinguals and tested in Spanish. In line with prior studies in this population and the sociolinguistic context of Barcelona [25,26], this classification represents a coarse, ecologically grounded proxy of bilingual language use/exposure rather than a comprehensive assessment of bilingual experience, which is unavailable for this historical cohort.

Our sample comprised 280 active bilinguals (166 female, mean age = 76 ± 4.69 years) and 287 passive bilinguals (180 female, mean age = 75.5 ± 4.48 years). All participants were recruited from Barcelona and spoke Castilian Spanish. Individuals whose first language was not Spanish and/or Catalan, as well as those from other regions of Spain or other countries, were excluded to minimize variability related to dialectal/language varieties and sociocultural backgrounds. This approach resulted in a relatively homogeneous sample and reduced the confounding effects of significant sociocultural differences and cross-country immigration. All procedures were approved by the ethics committee of Hospital Sant Pau and conducted in accordance with the Declaration of Helsinki. Participants or their legally authorized representative provided written informed consent.

### 2.2. Neuropsychological evaluation

Participants completed a comprehensive neuropsychological assessment (see details in [14]), including global cognition (via Mini–Mental State Examination, MMSE), verbal memory (Free and Cued Selective Reminding Test, FCSRT, immediate and delayed free and cued recall), visual memory (geometric figures recall from the Consortium to Establish a Registry for Alzheimer’s Disease, CERAD), attention/executive functions (EF; Trail-Making Test, TMT, Parts A and B, and Wechsler Adult Intelligence Scale digit span backwards), language (phonemic and semantic fluency tasks and Boston Naming Test, BNT), visuospatial/visuomotor/visuoperceptive functioning (CERAD geometric figures copy subtest, Visual Object Space and Perception, VOSP, number location subtest, and Poppelreuter overlapping figures test), and neuropsychiatric symptoms (neuropsychiatric inventory and Geriatric Depression Scale). For BNT, the percentage of correctly named objects was used to accommodate the use of the short vs. full versions across participants.

### 2.3. CSF and blood collection and analysis

CSF was obtained by lumbar puncture and collected in polypropylene tubes following international recommendations [27]. Core AD biomarkers (Aβ-42, Aβ-40, phosphorylated tau or p-tau 181, and t-tau) were measured in the fully-automated platform Lumipulse using commercially available kits (Fujirebio Europe, Ghent, Belgium) [24]. NfL and Chitinase-3-like protein 1 (YKL-40) were measured by enzyme-linked immunosorbent assay (ELISA), and GFAP by single molecule array (Simoa) [28,29]. We used three CSF measures, Aβ-42, Aβ-42/40 ratio, and p-tau 181, to confirm participants’ A+T+ stage.

Blood was collected at the time of lumbar puncture, and plasma was aliquoted in polypropylene tubes. Plasma p-tau 181 and 217 samples were measured in the Lumipulse fully-automated platform with commercially available kits [30]. NfL and GFAP were measured using Simoa [31]. In the biomarker analysis reported below, we included CSF and plasma measures that reflect AD-specific pathophysiology (amyloidosis: Aβ-42, Aβ-40, and Aβ-42/40 ratio; tau accumulation: p-tau and t-tau), non-specific neurodegeneration (NfL), and neuroinflammation (GFAP and YKL-40). Aβ, t-tau, and YKL-40 were represented in CSF only.

### 2.4. Statistical modeling and analysis

All statistical analyses were performed in *R* (v4.4.2) [32]. We treated neuropsychological measures as count data; dependent on variable dispersion and proportion of excessive zeros, we built Poisson or negative binomial generalized linear models (GLMs; via the “MASS” package, v7.3-65 [33]) or zero-inflated negative binomial models (via the “pscl” package, v1.5.9 [34]). For biofluid measures, we treated them as continuous data and built Gamma GLMs. Across these analyses, we evaluated the same effects of interest, including bilingual status alongside its interaction with education or sex. Regarding covariates, we controlled for age in the neuropsychological analyses, and controlled for age, MMSE (a proxy of overall cognitive impairment), and apolipoprotein E4 (*APOE*-4) status (considered for Aβ-related measures) in the biomarker analyses. We analyzed the effects of bilingualism and relevant interactions by comparing models via the likelihood ratio test, for which effect size was reported as Cohen’s w and exponentiated coefficients reported as rate ratios. Analyses were conducted separately for each neuropsychological and biomarker variable. Marginal means were estimated (via the “emmeans” package, v1.11.1 [35]) and post hoc pairwise comparisons were corrected using the Bonferroni procedure, for which effect size was expressed as incidence rate ratios (IRRs).

Based on the significant associations observed above, we conducted mediation analyses (via the “mediation” package, v4.5.0 [36]) to explore whether biomarkers mediated the association between bilingualism and cognition. We re-assessed associations between bilingualism and neuropsychological variables in participants with available biomarker data; only associations that remained significant were entered into subsequent mediation analyses. Specifically, we fitted a mediator model where the target biomarker was modeled as a function of bilingual status, and an outcome model where the retained neuropsychological variable was modeled as a function of bilingual status and the biomarker. Both models were implemented via GLMs and included the same covariates as the primary analyses. We computed the average causal mediation effect (ACME; indirect effect), average direct effect (ADE), and total effect, with confidence intervals (CIs) calculated from 1000 bootstrap samples.

## 3. Results

### 3.1. Demographic and clinical characteristics

The two language groups had comparable age and sex distribution (Table 1). However, active bilinguals had significantly more years of education than passive bilinguals (*p* < .001), which was accounted for in the subsequent modeling analyses. Additionally, groups did not differ in the proportion of participants classified as MCI vs. AD dementia (based on Global Deterioration Scale criteria) or in *APOE*-4 carrier distribution.

**Table 1.**
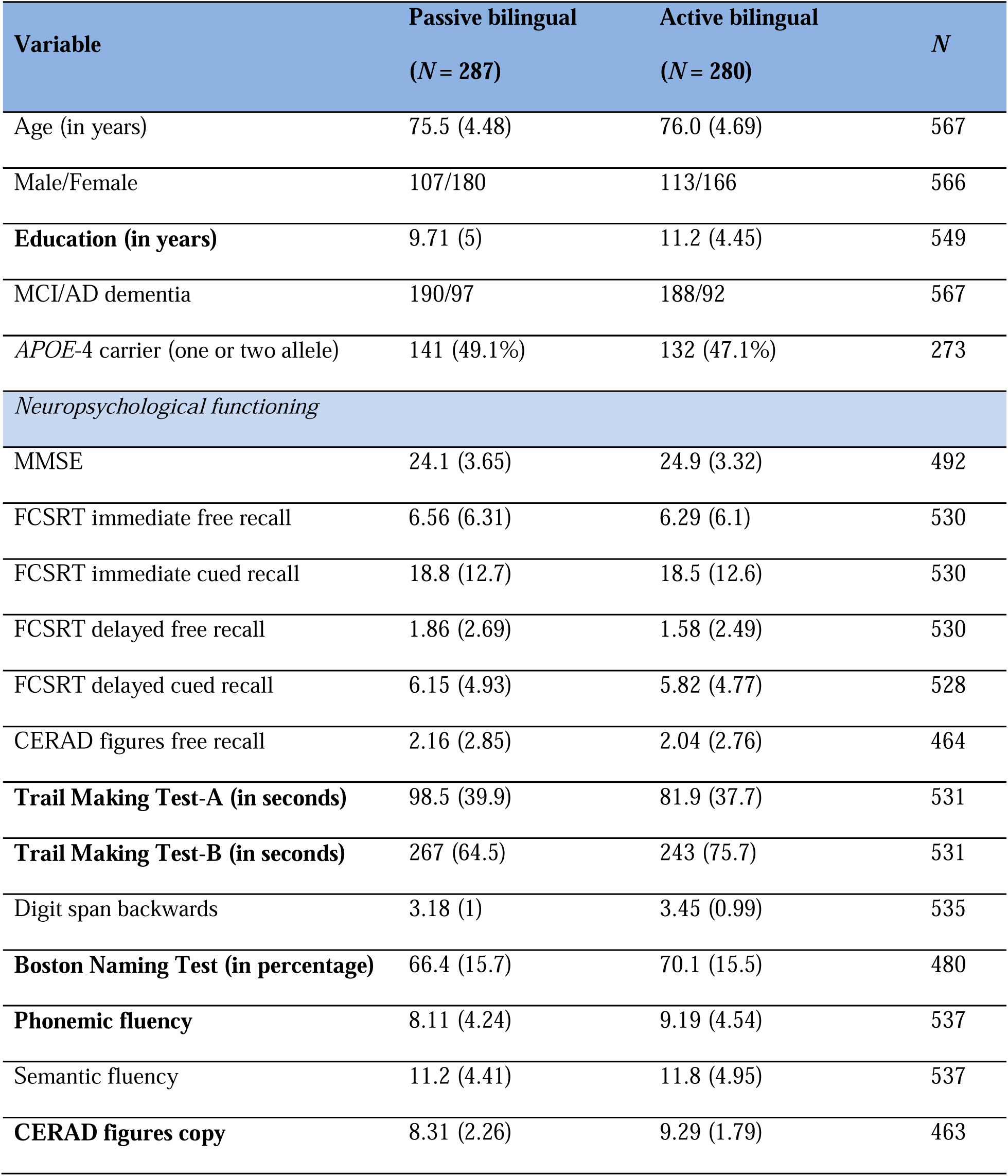
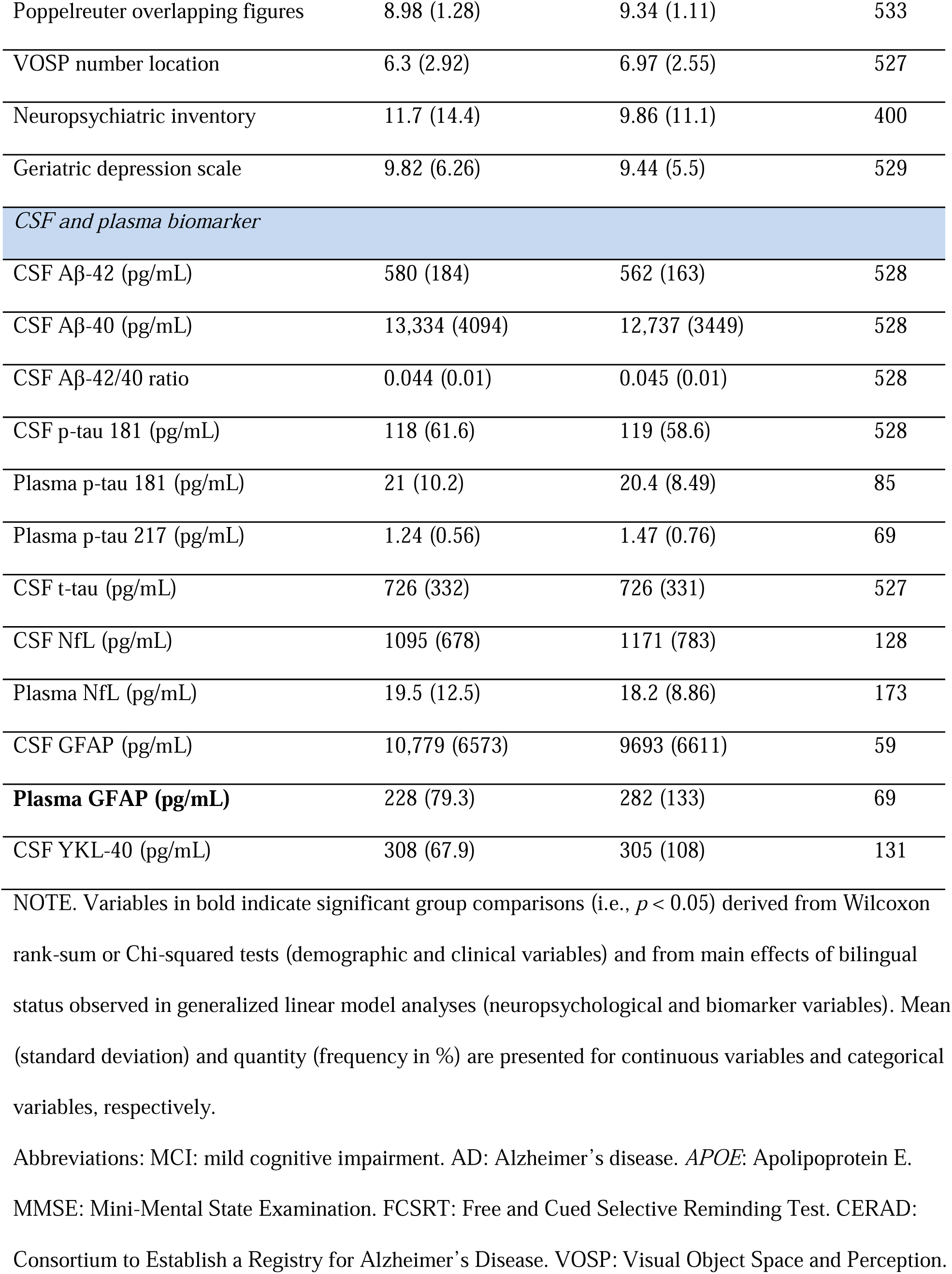
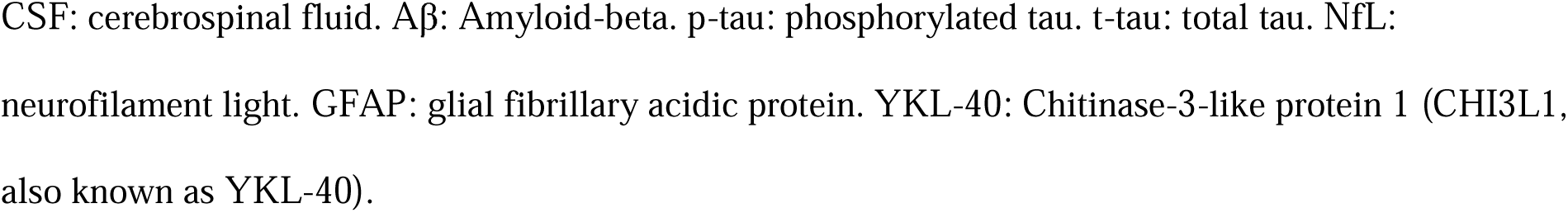
Descriptive statistics of demographic, clinical, neuropsychological, and biomarker data across.

| **Variable** | **Passive bilingual**<br>**(N = 287)** | **Active bilingual**<br>**(N = 280)** | **N** |
| --- | --- | --- | --- |
| Age (in years) | 75.5 (4.48) | 76.0 (4.69) | 567 |
| Male/Female | 107/180 | 113/166 | 566 |
| **Education (in years)** | 9.71 (5) | 11.2 (4.45) | 549 |
| MCI/AD dementia | 190/97 | 188/92 | 567 |
| *APOE-4* carrier (one or two allele) | 141 (49.1%) | 132 (47.1%) | 273 |
| *Neuropsychological functioning* |  |  |  |
| MMSE | 24.1 (3.65) | 24.9 (3.32) | 492 |
| FCSRT immediate free recall | 6.56 (6.31) | 6.29 (6.1) | 530 |
| FCSRT immediate cued recall | 18.8 (12.7) | 18.5 (12.6) | 530 |
| FCSRT delayed free recall | 1.86 (2.69) | 1.58 (2.49) | 530 |
| FCSRT delayed cued recall | 6.15 (4.93) | 5.82 (4.77) | 528 |
| CERAD figures free recall | 2.16 (2.85) | 2.04 (2.76) | 464 |
| **Trail Making Test-A (in seconds)** | 98.5 (39.9) | 81.9 (37.7) | 531 |
| **Trail Making Test-B (in seconds)** | 267 (64.5) | 243 (75.7) | 531 |
| Digit span backwards | 3.18 (1) | 3.45 (0.99) | 535 |
| **Boston Naming Test (in percentage)** | 66.4 (15.7) | 70.1 (15.5) | 480 |
| **Phonemic fluency** | 8.11 (4.24) | 9.19 (4.54) | 537 |
| Semantic fluency | 11.2 (4.41) | 11.8 (4.95) | 537 |
| **CERAD figures copy** | 8.31 (2.26) | 9.29 (1.79) | 463 |
| Poppelreuter overlapping figures | 8.98 (1.28) | 9.34 (1.11) | 533 |
| VOSP number location | 6.3 (2.92) | 6.97 (2.55) | 527 |
| Neuropsychiatric inventory | 11.7 (14.4) | 9.86 (11.1) | 400 |
| Geriatric depression scale | 9.82 (6.26) | 9.44 (5.5) | 529 |
| *CSF and plasma biomarker* |  |  |  |
| CSF Aβ-42 (pg/mL) | 580 (184) | 562 (163) | 528 |
| CSF Aβ-40 (pg/mL) | 13,334 (4094) | 12,737 (3449) | 528 |
| CSF Aβ-42/40 ratio | 0.044 (0.01) | 0.045 (0.01) | 528 |
| CSF p-tau 181 (pg/mL) | 118 (61.6) | 119 (58.6) | 528 |
| Plasma p-tau 181 (pg/mL) | 21 (10.2) | 20.4 (8.49) | 85 |
| Plasma p-tau 217 (pg/mL) | 1.24 (0.56) | 1.47 (0.76) | 69 |
| CSF t-tau (pg/mL) | 726 (332) | 726 (331) | 527 |
| CSF NfL (pg/mL) | 1095 (678) | 1171 (783) | 128 |
| Plasma NfL (pg/mL) | 19.5 (12.5) | 18.2 (8.86) | 173 |
| CSF GFAP (pg/mL) | 10,779 (6573) | 9693 (6611) | 59 |
| **Plasma GFAP (pg/mL)** | 228 (79.3) | 282 (133) | 69 |
| CSF YKL-40 (pg/mL) | 308 (67.9) | 305 (108) | 131 |

NOTE. Variables in bold indicate significant group comparisons (i.e., $p < 0.05$ ) derived from Wilcoxon rank-sum or Chi-squared tests (demographic and clinical variables) and from main effects of bilingual status observed in generalized linear model analyses (neuropsychological and biomarker variables). Mean (standard deviation) and quantity (frequency in %) are presented for continuous variables and categorical variables, respectively.
Abbreviations: MCI: mild cognitive impairment. AD: Alzheimer's disease. *APOE*: Apolipoprotein E. MMSE: Mini-Mental State Examination. FCSRT: Free and Cued Selective Reminding Test. CERAD: Consortium to Establish a Registry for Alzheimer's Disease. VOSP: Visual Object Space and Perception.
CSF: cerebrospinal fluid. Aβ : Amyloid-beta. p-tau: phosphorylated tau. t-tau: total tau. NfL: neurofilament light. GFAP: glial fibrillary acidic protein. YKL-40: Chitinase-3-like protein 1 (CHI3L1, also known as YKL-40).

### 3.2. Neuropsychological functioning

Controlling for age, sex, and education, we did not observe significant bilingualism effects on overall cognitive status (indexed by MMSE). However, accounting for the same covariates, active bilingualism was significantly associated with better performance on various neuropsychological outcomes across cognitive domains. For attention/EF, model comparisons revealed significant bilingual status effects on TMT-A (*p* < .001, Cohen’s w = 0.17) and TMT-B (*p* = .007, Cohen’s w = 0.12), with significantly faster responses in active than passive bilinguals (Figure 1A-B; Supplemental Tables 1-2). For language, we also found significant bilingual status effects on BNT (*p* = .04, Cohen’s w = 0.09) and phonemic fluency (*p* = .045, Cohen’s w = 0.09), with active bilinguals producing significantly more accurate responses or more items (Figure 1C-D; Supplemental Tables 3-4). As to visuospatial/visuomotor functioning, we first observed significant bilingual status effects on CERAD figures copy (*p* = .004, Cohen’s w = 0.13), in which active bilinguals outperformed passive bilinguals (Figure 1E; Supplemental Table 5). We also found a significant bilingual status × sex interaction predicting VOSP number location performance (*p* = .032, Cohen’s w = 0.09). Pairwise comparisons based on the optimal model (Supplemental Table 6) showed that active bilinguals had significantly higher scores than passive bilinguals within women only (adjusted *p* = .01, IRR = 0.88; Figure 1F), and men outperformed women across language groups (passive bilingual: adjusted *p* < .001, IRR = 1.3; active bilingual: adjusted *p* = .034, IRR = 1.12). There were no significant bilingualism effects on other neuropsychological variables (Supplemental Table 7), including no instances where active bilingualism was associated with worse performance of any neuropsychological measures.

**Figure 1.**
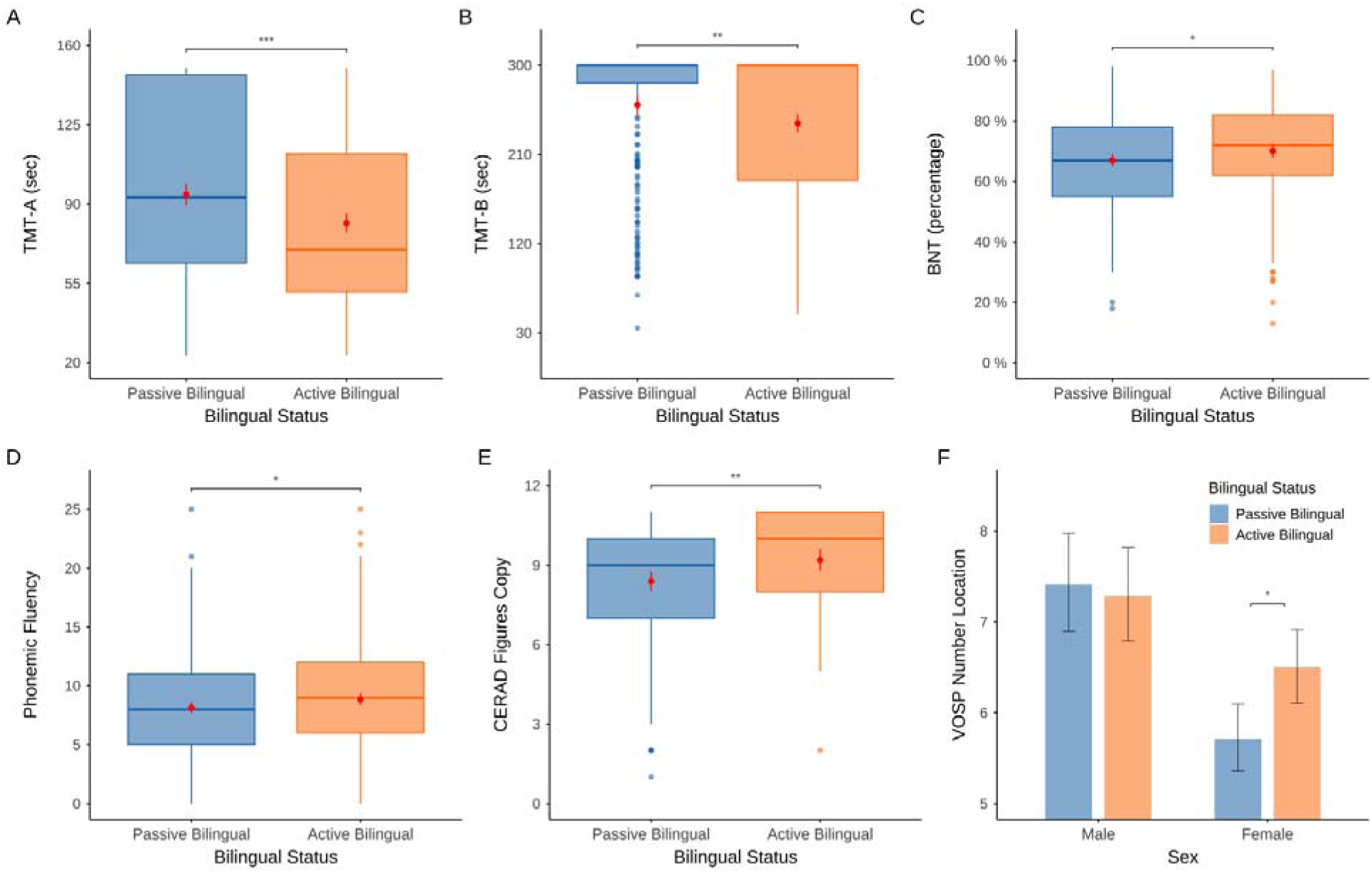
**Significant effects of bilingual status or its interaction with sex on neuropsychological functioning outcomes** NOTE. Active bilinguals outperformed passive bilinguals across measures, which was specific to female participants for the VOSP number location, only (Panel F). Asterisks indicate significant group comparisons after correction: *** *p* < .001, ** *p* < .01, * *p* < .05. The red dots and vertical lines in box plots for significant main effects (Panels A through E) represent predicted means and 95% confidence intervals computed from the optimal model, respectively. The error bars in the bar graphs for the significant interaction (Panel F) represent 95% confidence interval of the estimated means. Abbreviations: TMT: Trail Making Test. BNT: Boston Naming Test. CERAD: Consortium to Establish a Registry for Alzheimer’s Disease. VOSP: Visual Object Space and Perception.

### 3.3. CSF and plasma biomarkers

Controlling for age and cognitive impairment, we found multiple associations indicative of a “reduced pathological” biomarker profile in active bilinguals and therefore of resistance to AD-specific pathophysiologic processes and neuroinflammation. First, we found a significant bilingual status × sex interaction on CSF Aβ-42/40 ratio (*p* = .011, Cohen’s w = 0.02). Pairwise comparisons based on the optimal model (Supplemental Table 8) revealed a significantly higher Aβ-42/40 ratio—indicating reduced amyloid burden—in active compared to passive bilinguals in men (adjusted *p* = .044, rate ratio = 0.94; Figure 2A). For neuroinflammation biomarkers, we found a significant bilingual status × sex interaction on CSF YKL-40 (*p* = .019, Cohen’s w = 0.05; Supplemental Table 9), revealing that active bilingual women exhibited significantly lower CSF YKL-40 levels, indicative of decreased neuroinflammation, than passive bilingual women (adjusted *p* = .018, rate ratio = 1.15; Figure 2B). Lastly, we observed a significant bilingual status × education interaction on CSF GFAP (*p* = .014, Cohen’s w = 0.21; Supplemental Table 10), with lower levels of CSF GFAP in active bilinguals being associated with higher educational attainment (Figure 2C).

**Figure 2.**
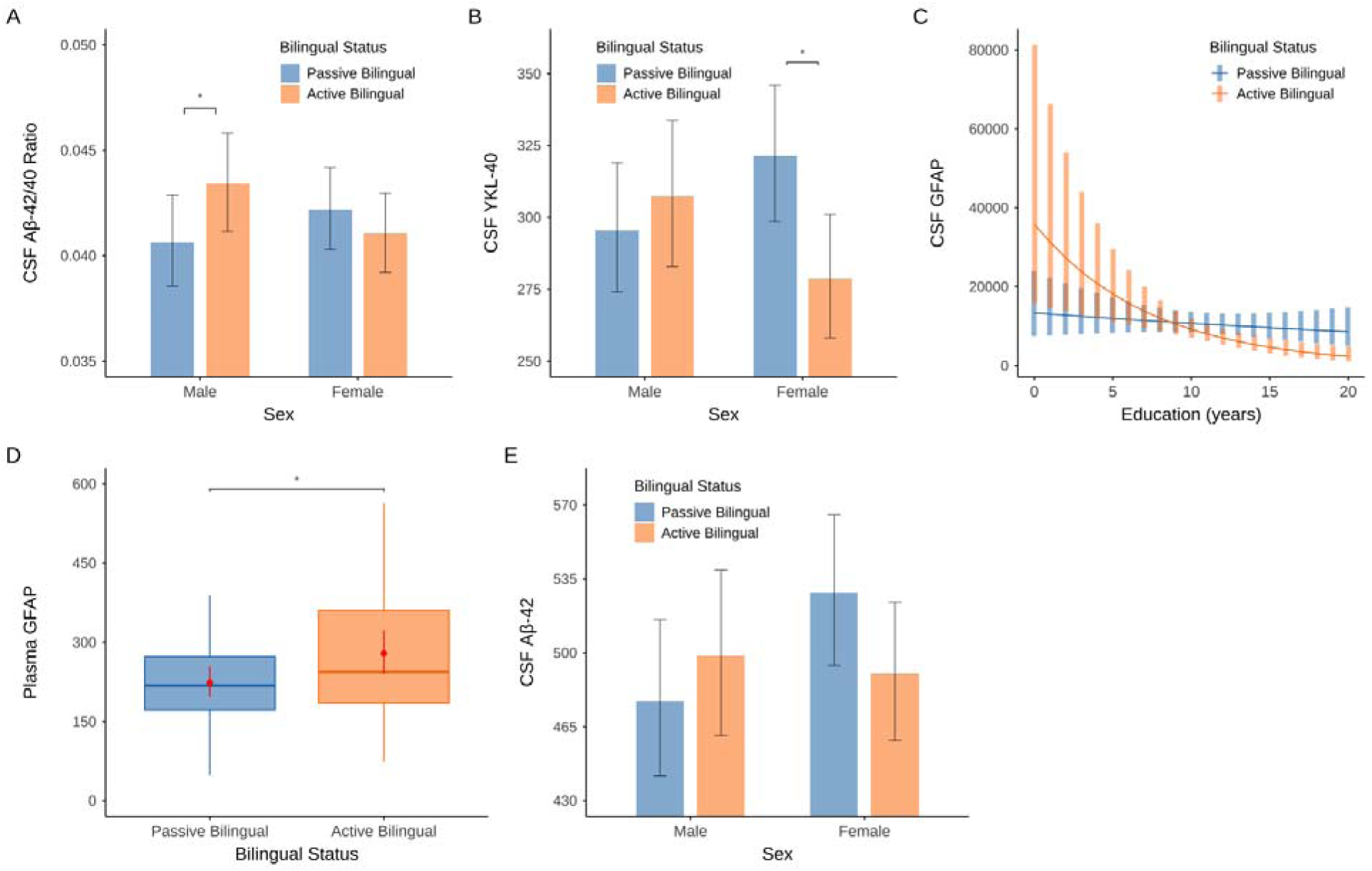
**Significant effects of bilingual status alongside its interaction with sex or education on biomarker levels** NOTE. Active bilinguals exhibited more favorable profiles in CSF Aβ-42/40 ratio (male participants; Panel A), CSF YKL-40 (female participants; B), and CSF GFAP (participants with higher education; C), as well as less favorable profiles in plasma GFAP (D) and CSF Aβ-42 (female participants; E). Asterisks indicate significant group comparisons after correction: *** *p* < .001, ** *p* < .01, * *p* < .05. The error bars in the bar graphs (Panels A, B, and E) and line graphs (Panel C) for significant interactions represent 95% confidence interval of the estimated means. The red dots and vertical lines in box plots (Panel D) for significant main effects represent predicted means and 95% confidence intervals computed from the optimal model, respectively. Abbreviations: CSF: cerebrospinal fluid. Aβ: Amyloid-beta. YKL-40: Chitinase-3-like protein 1 (CHI3L1, also known as YKL-40). GFAP: glial fibrillary acidic protein.

We also found two associations indicative of an “increased pathological” biomarker profile in active bilinguals, while controlling for age and cognitive impairment, and therefore provide evidence of resilience. First, we observed a significant bilingual status effect on plasma GFAP (*p* = .024, Cohen’s w = 0.11; Supplemental Table 11), where active bilinguals had significantly higher levels than passive bilinguals (Figure 2D). In addition, there was a significant bilingual status × sex interaction on CSF Aβ-42 (*p* = .029, Cohen’s w = 0.03; Supplemental Table 12). Among passive bilinguals, men had significantly lower Aβ-42 levels than women (adjusted *p* = .018, rate ratio = 0.9; Figure 2E). We did not find significant bilingualism effects on other CSF and plasma biomarkers (Supplemental Table 13).

### 3.4. Mediation analysis of bilingualism’s protective associations with neuropsychological functioning

Considering only the “protective” associations that showed more favorable profiles among active bilinguals identified above, we focused on the three CSF biomarkers: Aβ-42/40 ratio, YKL-40, and GFAP. Based on the subset of participants with data for these biomarkers, we re-evaluated the associations between bilingual status and neuropsychological outcomes (Table 2), eight of which remained significant and were used in the mediation analyses. However, none of the CSF biomarkers significantly mediated the relationship between active bilingualism and improved cognitive performance across domains (Figure 3): attention/EF (TMT-A and TMT-B), language (phonemic fluency), and visuospatial/visuomotor functioning (CERAD figures copy).

**Figure 3.**
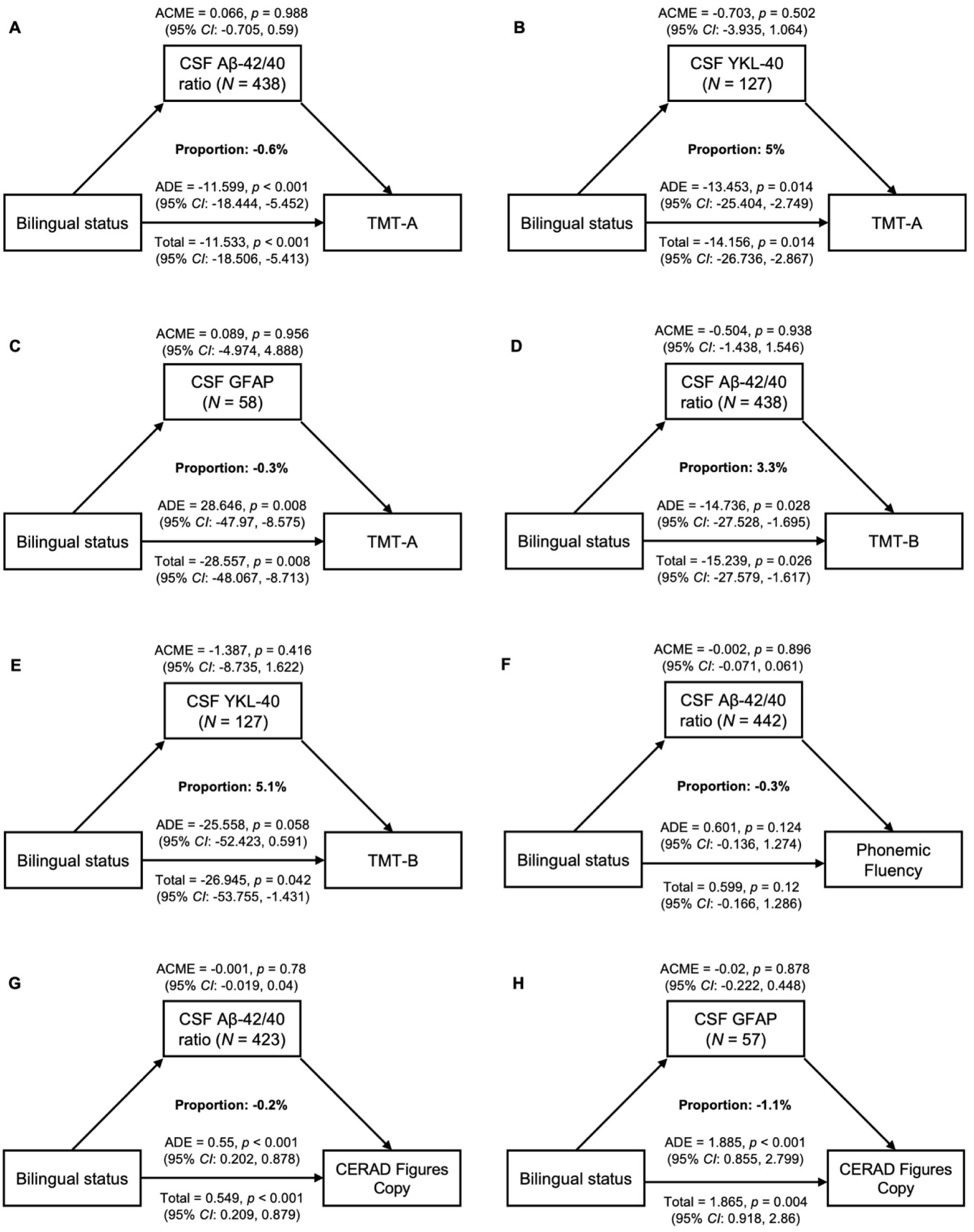
**Diagrams illustrating the mediation effects of biofluid markers on the relationship between bilingual status and neuropsychological functioning** NOTE. The mediation or indirect effect (ACME), direct effect (ADE), and total effect are reported. The *p*-value and 95% CI were estimated using bootstrapping samples. No significant mediation effects were found across measures. *N* represents the number of participants with data of the biomarker, neuropsychological measure, and covariates (i.e., education, sex, age, and MMSE). The mediator model included the treatment and mediator, whereas the outcome model also included the outcome. Abbreviations: ACME: average causal mediation effect. ADE: average direct effect. CSF: cerebrospinal fluid. YKL-40: Chitinase-3-like protein 1 (CHI3L1). TMT: Trail Making Test. Aβ: Amyloid-beta. GFAP: glial fibrillary acidic protein. CERAD: Consortium to Establish a Registry for Alzheimer’s Disease.

**Table 2.**
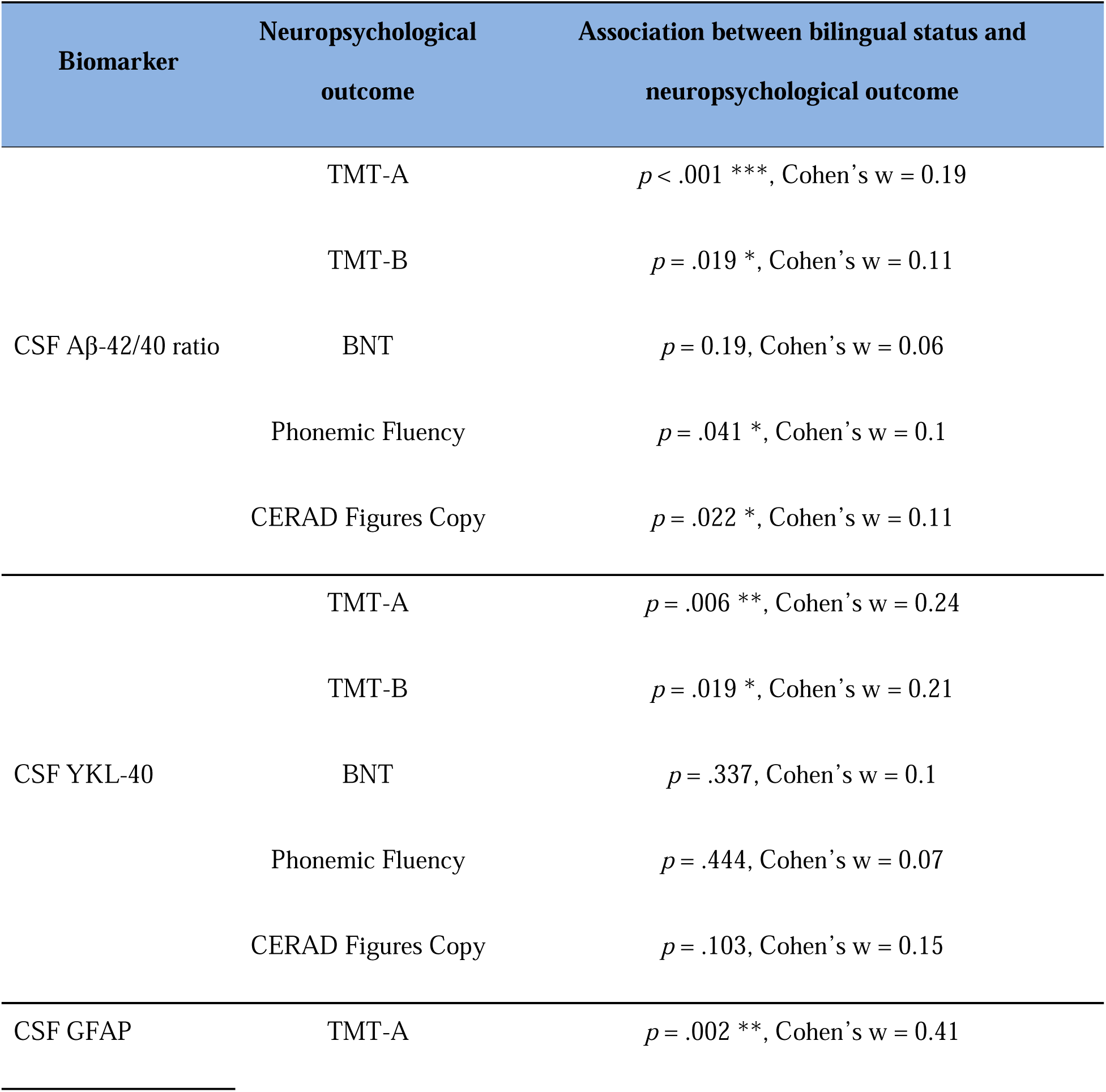
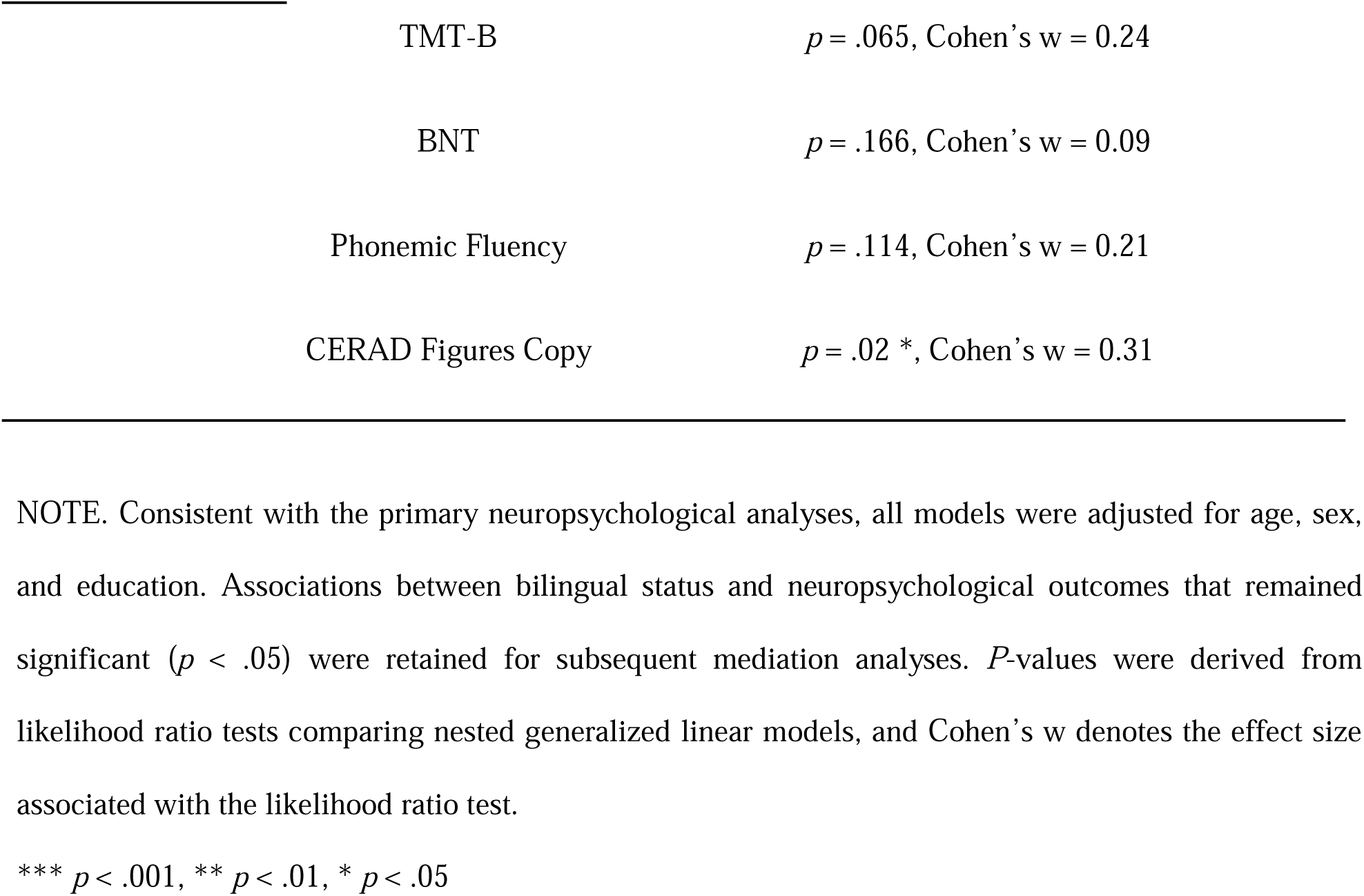
Associations between bilingual status and neuropsychological outcomes in the subset of participants with biomarker data showing favorable profiles in active bilinguals.

## 4. Discussion

In this study, we investigated the impact of bilingualism on neuropsychological and biomarker profiles in an AD cohort at the same biological stage (A+T+). We found that active bilingualism was linked to better neuropsychological performance across certain but not all cognitive domains (attention/EF, language, visuospatial/visuomotor), demonstrating resilience to neurodegenerative pathophysiology. Further, it was associated with more favorable CSF biomarker profiles (Aβ-42/40 ratio, YKL-40, GFAP) when accounting for interactions with sex or education and controlling for age and cognitive impairment, indicating resistance to pathological processes. These findings suggest that bilingualism may protect against cognitive decline through both resilience and resistance mechanisms.

### 4.1. Effects of bilingualism on neuropsychological functioning

We observed that independent of age and education, active bilinguals outperformed passive bilinguals on tests assessing attention/EF (TMT-A, TMT-B), language (BNT, phonemic fluency), and visuospatial/visuomotor functioning (CERAD figures copy and VOSP number location, the latter in women only). This provides evidence for resilience [16–18], given comparable age, AD biological stage, and clinical stage between groups. Yet, there have been mixed findings about these cognitive measures in the literature, likely due to variability in sample characteristics, disease stage, and control of confounders.

For EF, prior studies in clinically diagnosed AD individuals often report no differences in TMT performance between language groups, for example, when comparing English monolinguals with Welsh-English bilinguals [37] or with English bilinguals from diverse language backgrounds [38]. Results in the healthy aging population are inconsistent: a bilingual advantage was found in Swiss participants (TMT-A; though only when education and occupation were not controlled) [39] and in Spanish participants (TMT-B in late bilinguals vs. monolinguals, accounting for multiple confounders) [12], whereas a monolingual advantage was observed in French speakers (smaller B-A difference in response times relative to French-Italian bilinguals) [40]. However, focusing on healthy individuals and those with MCI or AD from the same Spanish-Catalan bilingual population studied here, Calabria et al [25] found that active bilingualism was associated with better conflict resolution in a spatial Stroop task, which aligns with our finding about enhanced EF in active bilinguals.

Regarding language, our results lend further support to the link between bilingualism and better phonemic fluency, consistent with prior work in healthy bilinguals from the Spanish-Catalan population [41] and from other sociolinguistic contexts across countries [3,40,42,43]. This pattern likely reflects EF processes underpinning phonemic fluency [44], and our study extends it to pathological aging. Yet, some AD studies without biomarker confirmation found no group differences in letter fluency [37] or lower scores in bilinguals [38]. In comparison, a bilingual disadvantage in semantic fluency and BNT is often reported, possibly due to cross-language interference and reduced vocabulary in bilinguals [43]. However, these studies usually test the less dominant language and focus on healthy younger participants [44,45], differing from our sample and others documenting more favorable performance in bilinguals.

For visuospatial/visuomotor skills, we observed superior performance on the CERAD figures copy test in active bilinguals. This aligns with Perani et al [46], who showed better visuospatial short-term memory in German-Italian bilinguals with early-stage AD compared with Italian monolinguals. Similar findings have been reported in healthy bilinguals. For example, Estanga et al [12] found that both early and late bilinguals outperformed monolinguals on the Judgement of Line Orientation Test. McLeay [47] found faster mental rotation performance in Welsh-English bilinguals than in monolinguals. We also replicated the well-documented sex difference in visuospatial ability [10,48], with men outperforming women on VOSP number location across language groups. Notably, the bilingual benefit was evident only in women, which echoes prior findings that bilingualism may modulate sex differences in cognitive performance [21,22].

### 4.2. Effects of bilingualism on biofluid markers of brain health

Our analyses revealed multiple interactions between bilingualism and sex or education on Aβ deposition and neuroinflammation biomarkers. Active bilinguals exhibited a more favorable CSF profile—indicating less pathology and thus resistance [16–18]—with a higher Aβ-42/40 ratio in men, lower YKL-40 in women, and lower GFAP in individuals with higher education, when compared to passive bilinguals. No bilingualism effects emerged for CSF tau or neurodegeneration markers (p-tau, t-tau, and NfL). Our results differ from those of Estanga et al [12], who studied cognitively healthy adults (aged 40–80 years) and found lower CSF t-tau levels in early bilinguals relative to monolinguals.

Active bilinguals also showed greater pathology—demonstrating resilience [16–18]—with higher plasma GFAP across sexes. Although we did not examine education effects independently on Aβ deposition, prior PET evidence has linked higher education to increased Aβ accumulation in AD individuals matched for cognitive severity [9], suggesting that education and bilingualism may both confer resilience. Interestingly, CSF and plasma GFAP were associated with different “protective” effects (resilience-resistance) while putatively measuring the same pathophysiologic process. We suspect this may reflect temporal dynamics of AD pathophysiology, as plasma GFAP levels are higher than those of CSF in preclinical and symptomatic stages [49].

### 4.3. Mediation effects of biomarkers on the relationship between bilingualism and neuropsychological functioning

We explored whether the positive bilingualism-cognition link was mediated by markers of various pathophysiologic mechanisms involved in neurodegeneration but did not find significant mediation effects. This finding should not be interpreted as evidence against a biological contribution to bilingualism-related cognitive differences. It is possible that unmeasured physiologic mechanisms reflected by markers of synaptic function or unmeasured pathways such as those between biofluid and structural brain measures may underlie the observed bilingual cognitive advantages. Estanga et al [12] investigated whether bilingualism moderated CSF biomarker-cognition relations in healthy older adults and found no significant effects either. Despite these null results, bilingualism, as an environmental enrichment form, has been postulated to enhance neurogenesis and synaptogenesis via multiple neural factors [50] and may represent a cost-effective approach to maintaining brain health.

### 4.4. Limitations and future directions

Constrained by data availability in this historical cohort, we operationalized bilingualism by comparing active vs. passive bilinguals, which did not capture participants’ multidimensional bilingual experience. However, meaningful comparisons along the mono-bilingualism continuum depend on sociocultural contexts; our findings remain relevant to passive bilingual communities where exposure to additional languages does not necessarily imply speaking proficiency. Moreover, the cross-sectional design of our study limits inferences about the temporal dynamics underlying resilience and resistance, particularly with respect to whether bilingualism is associated with trajectories of biomarker change over time or reflects interindividual differences at disease onset.

Future research should examine more nuanced bilingual characteristics (e.g., language dominance, proficiency), which may uncover additional relations of interest. For instance, incorporating age of acquisition might identify subgroup-specific effects (e.g., early vs. late bilingual differences [12]). Further, accounting for more fine-grained socioeconomic indicators (e.g., income, occupation) in future investigations may help disentangle bilingualism-related effects from broader life-course factors, while longitudinal designs will be necessary to more directly examine the causal pathways linking bilingualism to cognitive and biomarker outcomes.

### 4.5. Conclusion

This study demonstrates the protective effects of active bilingualism in older adults with biomarker-confirmed AD. Our results lend further support to bilinguals’ better cognitive performance across domains (attention/EF, language, and visuospatial/visuomotor), highlighting cognitive resilience. At the biological level, we provide novel evidence of a more favorable CSF biomarker profile in active bilinguals—a higher Aβ-42/40 ratio and lower levels of neuroinflammation markers (YKL-40 and GFAP)—thereby indicating resistance. Overall, these findings suggest that bilingual experience may confer benefits through both resilience and resistance to AD pathology mechanisms.

## Data Availability

All data produced in the present study are available upon reasonable request to the authors.

## Acknowledgements

The authors would like to thank all participants for their contribution to the research.

## References

[1] Celik S, Kokje E, Meyer P, Frölich L, Teichmann B. Does bilingualism influence neuropsychological test performance in older adults? A systematic review. Appl Neuropsychol Adult. 2020;29(4):855–873. doi:10.1080/23279095.2020.1788032

[2] de Leon J, Grasso SM, Welch A, et al. Effects of bilingualism on age at onset in two clinical Alzheimer’s disease variants. Alzheimers Dement. 2020;16(12):1704–1713. doi:10.1002/alz.12170

[3] Venugopal A, Paplikar A, Varghese FA, et al. Protective effect of bilingualism on aging, MCI, and dementia: A community-based study. Alzheimers Dement. 2024;20(4):2620–2631. doi:10.1002/alz.13702

[4] Prior A, Macwhinney B. A bilingual advantage in task switching. Biling (Camb Engl). 2010;13(2):253–262. doi:10.1017/S1366728909990526

[5] Abutalebi J, Canini M, Della Rosa PA, Sheung LP, Green DW, Weekes BS. Bilingualism protects anterior temporal lobe integrity in aging. Neurobiol Aging. 2014;35(9):2126–2133. doi:10.1016/j.neurobiolaging.2014.03.010

[6] van den Noort M, Vermeire K, Bosch P, et al. A systematic review on the possible relationship between bilingualism, cognitive decline, and the onset of dementia. Behavioral Sciences. 2019;9(7). doi:10.3390/bs9070081

[7] Beach TG, Monsell SE, Phillips LE, Kukull W. Accuracy of the clinical diagnosis of Alzheimer disease at National Institute on Aging Alzheimer Disease Centers, 2005-2010. J Neuropathol Exp Neurol. 2012;71(4):266–273. doi:10.1097/NEN.0b013e31824b211b

[8] Livingston G, Huntley J, Liu KY, et al. Dementia prevention, intervention, and care: 2024 report of the Lancet standing Commission. Lancet. 2024;404(10452):572–628. doi:10.1016/S0140-6736(24)01296-0

[9] Kemppainen NM, Aalto S, Karrasch M, et al. Cognitive reserve hypothesis: Pittsburgh Compound B and fluorodeoxyglucose positron emission tomography in relation to education in mild Alzheimer’s disease. Ann Neurol. 2008;63(1):112–118. doi:10.1002/ana.21212

[10] Weiss EM, Kemmler G, Deisenhammer EA, Fleischhacker WWolfgang, Delazer M. Sex differences in cognitive functions. Pers Individ Dif. 2003;35(4):863–875. doi:10.1016/s0191-8869(02)00288-x

[11] Bridel C, van Wieringen WN, Zetterberg H, et al. Diagnostic value of cerebrospinal fluid neurofilament light protein in neurology: A systematic review and meta-analysis. JAMA Neurol. 2019;76(9):1035–1048. doi:10.1001/jamaneurol.2019.1534

[12] Estanga A, Ecay-Torres M, Ibañez A, et al. Beneficial effect of bilingualism on Alzheimer’s disease CSF biomarkers and cognition. Neurobiol Aging. 2017;50:144–151. doi:10.1016/j.neurobiolaging.2016.10.013

[13] Grasso SM, Clark AL, Petersen M, O’Bryant S; Health and Aging Brain Study (HABS HD) Study Team. Bilingual neurocognitive resiliency, vulnerability, and Alzheimer’s disease biomarker correlates in Latino older adults enrolled in the Health and Aging Brain Study - Health Disparities (HABS-HD). Alzheimers Dement (Amst). 2023;15(4):e12509. doi:10.1002/dad2.12509

[14] Alcolea D, Clarimón J, Carmona-Iragui M, et al. The Sant Pau Initiative on Neurodegeneration (SPIN) cohort: A data set for biomarker discovery and validation in neurodegenerative disorders. Alzheimers Dement (N Y). 2019;5:597–609. doi:10.1016/j.trci.2019.09.005

[15] Jack CR Jr, Bennett DA, Blennow K, et al. NIA-AA Research Framework: Toward a biological definition of Alzheimer’s disease. Alzheimers Dement. 2018;14(4):535–562. doi:10.1016/j.jalz.2018.02.018

[16] Arenaza-Urquijo EM, Vemuri P. Resistance vs resilience to Alzheimer disease: Clarifying terminology for preclinical studies. Neurology. 2018;90(15):695–703. doi:10.1212/WNL.0000000000005303

[17] Arenaza-Urquijo EM, Vemuri P. Improving the resistance and resilience framework for aging and dementia studies. Alzheimers Res Ther. 2020;12(1):41. doi:10.1186/s13195-020-00609-2

[18] Montine TJ, Cholerton BA, Corrada MM, et al. Concepts for brain aging: Resistance, resilience, reserve, and compensation. Alzheimers Res Ther. 2019;11(1):22. doi:10.1186/s13195-019-0479-y

[19] Stern Y, Albert M, Barnes CA, Cabeza R, Pascual-Leone A, Rapp PR. A framework for concepts of reserve and resilience in aging. Neurobiol Aging. 2023;124:100–103. doi:10.1016/j.neurobiolaging.2022.10.015

[20] Cabeza R, Albert M, Belleville S, et al. Maintenance, reserve and compensation: The cognitive neuroscience of healthy ageing. Nat Rev Neurosci. 2018;19(11):701–710. doi:10.1038/s41583-018-0068-2

[21] Subramaniapillai S, Rajah MN, Pasvanis S, Titone D. Bilingual experience and executive control over the adult lifespan: The role of biological sex. Biling (Camb Engl). 2018;22(04):733–751. doi:10.1017/s1366728918000317

[22] Gonzalez-Marquez, M. Language, thought, and . . . brain? In: Spivey M, McRae K, Joanisse M, eds. The Cambridge Handbook of Psycholinguistics. Cambridge University Press; 2012:674–692. doi: 10.1017/CBO9781139029377.035

[23] Calvo N, Phillips N, Bialystok E, Einstein G. Biological sex and bilingualism: Its impact on risk and resilience for dementia. Alzheimers Dement (Amst). 2026;18(1):e70255. doi:10.1002/dad2.70255

[24] Alcolea D, Pegueroles J, Muñoz L, et al. Agreement of amyloid PET and CSF biomarkers for Alzheimer’s disease on Lumipulse. Ann Clin Transl Neurol. 2019;6(9):1815–24. doi:10.1002/acn3.50873

[25] Calabria M, Hernández M, Cattaneo G, et al. Active bilingualism delays the onset of mild cognitive impairment. Neuropsychologia. 2020;146:107528. doi: 10.1016/j.neuropsychologia.2020.107528

[26] Cattaneo G, Costa A, Gironell A, Calabria M. On the specificity of bilingual language control: A study with Parkinson’s disease patients. Biling (Camb Engl). 2019;23(3):570–578. doi: 10.1017/s136672891900004x

[27] Alcolea, D. et al. Feasibility of lumbar puncture in the study of cerebrospinal fluid biomarkers for Alzheimer’s disease: A multicenter study in Spain. J. Alzheimers Dis. 2014;39, 719–726. doi:10.3233/JAD-131334

[28] Illán-Gala, I. et al. CSF sAPPβ, YKL-40, and NfL along the ALS-FTD spectrum. Neurology. 2018;91(17), e1619–e1628. doi:10.1212/WNL.0000000000006383

[29] Montoliu-Gaya L, Alcolea D, Ashton NJ, et al. Plasma and cerebrospinal fluid glial fibrillary acidic protein levels in adults with Down Syndrome: A longitudinal cohort study. EBioMedicine. 2023;90:104547. doi:10.1016/j.ebiom.2023.104547

[30] Arranz J, Zhu N, Rubio-Guerra S, et al. Diagnostic performance of plasma pTau217, pTau181, Aβ1-42 and Aβ1-40 in the LUMIPULSE automated platform for the detection of Alzheimer disease. Alzheimers Res Ther. 2024;16(1):139. doi:10.1186/s13195-024-01513-9

[31] Alcolea D, Delaby C, Muñoz L, et al. Use of plasma biomarkers for AT(N) classification of neurodegenerative dementias. J Neurol Neurosurg Psychiatry. 2021;92(11):1206–1214. doi:10.1136/jnnp-2021-326603

[32] R Core Team. R: A Language and Environment for Statistical Computing. Vienna, Austria: R Foundation for Statistical Computing; 2024. https://www.R-project.org/

[33] Venables WN, Ripley BD. (2002). Modern Applied Statistics with S. 4th ed. Springer; 2002.

[34] Zeileis A, Kleiber C, Jackman S. Regression models for count data in R. J Stat Softw. 2008; 27(8):1–25. doi:10.18637/jss.v027.i08

[35] . Lenth R. emmeans: Estimated Marginal Means, aka Least-Squares Means. R package version 1.11.1-00001. 2025. https://rvlenth.github.io/emmeans/

[36] Tingley D, Yamamoto T, Hirose K, Keele L, Imai K. mediation: R package for causal mediation analysis. J Stat Softw. 2014;59(5):1–38. doi: 10.18637/jss.v059.i05

[37] Clare L, Whitaker CJ, Craik FI, et al. Bilingualism, executive control, and age at diagnosis among people with early-stage Alzheimer’s disease in Wales. J Neuropsychol. 2016;10(2):163–185. doi:10.1111/jnp.12061

[38] Anderson JA, Saleemi S, Bialystok E. Neuropsychological assessments of cognitive aging in monolingual and bilingual older adults. J Neurolinguistics. 2017;43(A):17–27. doi:10.1016/j.jneuroling.2016.08.001

[39] Ihle A, Oris M, Fagot D, Kliegel M. The relation of the number of languages spoken to performance in different cognitive abilities in old age. J Clin Exp Neuropsychol. 2016;38(10):1103–1114. doi:10.1080/13803395.2016.1197184

[40] Massa E, Köpke B, El Yagoubi R. Age-related effect on language control and executive control in bilingual and monolingual speakers: Behavioral and electrophysiological evidence. Neuropsychologia. 2020;138:107336. doi:10.1016/j.neuropsychologia.2020.107336

[41] Olabarrieta-Landa L, Benito-Sánchez I, Alegret M, et al. Letter verbal fluency in Spanish-, Basque-, and Catalan-speaking individuals: Does the selection of the letters influence the outcome? J Speech Lang Hear Res. 2019;62(7):2400–2410. doi:10.1044/2019_JSLHR-L-18-0365

[42] Friesen DC, Luo L, Luk G, Bialystok E. Proficiency and control in verbal fluency performance across the lifespan for monolinguals and bilinguals. Lang Cogn Neurosci. 2015;30(3):238–250. doi:10.1080/23273798.2014.918630

[43] Marsh JE, Hansson P, Sörman DE, Ljungberg JK. Executive processes underpin the bilingual advantage on phonemic fluency: Evidence from analyses of switching and clustering. Front Psychol. 2019;10:1355. doi:10.3389/fpsyg.2019.01355

[44] Roberts PM, Garcia LJ, Desrochers A, Hernandez D. English performance of proficient bilingual adults on the Boston Naming Test. Aphasiology. 2002;16(4-6):635–645. doi: 10.1080/02687030244000220

[45] Portocarrero JS, Burright RG, Donovick PJ. Vocabulary and verbal fluency of bilingual and monolingual college students. Arch Clin Neuropsychol. 2007;22(3):415–422. doi:10.1016/j.acn.2007.01.015

[46] Perani D, Farsad M, Ballarini T, et al. The impact of bilingualism on brain reserve and metabolic connectivity in Alzheimer’s dementia. Proc Natl Acad Sci U S A. 2017;114(7):1690–1695. doi:10.1073/pnas.1610909114

[47] McLeay H. The relationship between bilingualism and the performance of spatial tasks. Int J Biling Educ Biling. 2003;6(6):423–438. doi:10.1080/13670050308667795

[48] Resnick SM. Sex differences in mental rotations: An effect of time limits? Brain Cogn. 1993;21(1):71–79. doi:10.1006/brcg.1993.1005

[49] Benedet AL, Milà-Alomà M, Vrillon A, et al. Differences between plasma and cerebrospinal fluid glial fibrillary acidic protein levels across the Alzheimer disease continuum. JAMA Neurol. 2021;78(12):1471–1483. doi:10.1001/jamaneurol.2021.3671

[50] Kim S, Jeon SG, Nam Y, Kim HS, Yoo DH, Moon M. Bilingualism for dementia: Neurological mechanisms associated with functional and structural changes in the brain. Front Neurosci. 2019;13:1224. doi:10.3389/fnins.2019.01224

