## Supplemental Material for "Bilingualism’s protective effects in Alzheimer’s disease: Mechanisms of resilience and resistance"

Supplemental Table 1. Summary of the optimal model (negative binomial GLM) for *TMT-A*, predicted by the significant effect of bilingual status while accounting for education, sex, and age

| Model: glm.nb(formula = TMTA ~ BilingualStatus + Education + Sex + Age, link = log) | | | | | |
| --- | --- | --- | --- | --- | --- |
| Coefficients: |  |  |  |  |  |
|  | Estimate | Incidence Rate Ratio | Standard Error | z value | Pr (>\|z\|) |
| Intercept | 3.66067 | 38.9 | 0.31216 | 11.73 | < 0.001 *** |
| BilingualStatus: Active bilingual | -0.1425 | 0.867 | 0.03619 | -3.94 | **< 0.001** *** |
| Education | -0.0246 | 0.976 | 0.00389 | -6.33 | < 0.001 *** |
| Sex: Female | 0.12167 | 1.13 | 0.03732 | 3.26 | 0.001 ** |
| Age | 0.01428 | 1.01 | 0.00395 | 3.62 | < 0.001 *** |
| Null deviance: 651.03 on 529 degrees of freedom  Residual deviance: 543.23 on 525 degrees of freedom  (37 observations deleted due to missingness)  AIC: 5269 | | | | | |

*** *p* < 0.001, ** *p* < 0.01, * *p* < 0.05

Supplemental Table 2. Summary of the optimal model (negative binomial GLM) for *TMT-B*, predicted by the significant effect of bilingual status while accounting for education, sex, and age

| Model: glm.nb(formula = TMTB ~ BilingualStatus + Education + Sex + Age, link = log) | | | | | |
| --- | --- | --- | --- | --- | --- |
| Coefficients: |  |  |  |  |  |
|  | Estimate | Incidence Rate Ratio | Standard Error | z value | Pr (>\|z\|) |
| Intercept | 4.63668 | 103 | 0.23524 | 19.71 | < 0.001 *** |
| BilingualStatus: Active bilingual | -0.0732 | 0.929 | 0.02727 | -2.68 | **0.0073** ** |
| Education | -0.01861 | 0.982 | 0.00292 | -6.36 | < 0.001 *** |
| Sex: Female | 0.08348 | 1.09 | 0.02811 | 2.97 | 0.003 ** |
| Age | 0.01422 | 1.01 | 0.00297 | 4.78 | < 0.001 *** |
| Null deviance: 642.71 on 529 degrees of freedom  Residual deviance: 544.44 on 525 degrees of freedom  (37 observations deleted due to missingness)  AIC: 6107 | | | | | |

*** *p* < 0.001, ** *p* < 0.01, * *p* < 0.05

Supplemental Table 3. Summary of the optimal model (negative binomial GLM) for *BNT*, predicted by the significant effect of bilingual status while accounting for education, sex, and age

| Model: glm.nb(formula = BNT ~ BilingualStatus + Education + Sex + Age, link = log) | | | | | |
| --- | --- | --- | --- | --- | --- |
| Coefficients: |  |  |  |  |  |
|  | Estimate | Incidence Rate Ratio | Standard Error | z value | Pr (>\|z\|) |
| Intercept | 4.74925 | 115 | 0.1837 | 25.85 | < 0.001 *** |
| BilingualStatus: Active bilingual | 0.04422 | 1.05 | 0.02151 | 2.06 | **0.0398** * |
| Education | 0.00895 | 1.01 | 0.00232 | 3.86 | < 0.001 *** |
| Sex: Female | -0.06869 | 0.934 | 0.02211 | -3.11 | 0.002 ** |
| Age | -0.00796 | 0.992 | 0.00232 | -3.42 | < 0.001 *** |
| Null deviance: 553.21 on 479 degrees of freedom  Residual deviance: 501.15 on 475 degrees of freedom  (87 observations deleted due to missingness)  AIC: 4028 | | | | | |

*** *p* < 0.001, ** *p* < 0.01, * *p* < 0.05

Supplemental Table 4. Summary of the optimal model (negative binomial GLM) for *Phonemic Fluency*, predicted by the significant effect of bilingual status while accounting for education, sex, and age

| Model: glm.nb(formula = PhonemicFluency ~ BilingualStatus + Education + Sex + Age, link = log) | | | | | |
| --- | --- | --- | --- | --- | --- |
| Coefficients: |  |  |  |  |  |
|  | Estimate | Incidence Rate Ratio | Standard Error | z value | Pr (>\|z\|) |
| Intercept | 2.59523 | 13.4 | 0.36422 | 7.13 | < 0.001 *** |
| BilingualStatus: Active bilingual | 0.08449 | 1.09 | 0.04218 | 2 | **0.045** * |
| Education | 0.03449 | 1.04 | 0.00447 | 7.71 | < 0.001 *** |
| Sex: Female | 0.01061 | 1.01 | 0.04359 | 0.24 | 0.808 |
| Age | -0.01143 | 0.989 | 0.00461 | -2.48 | 0.013 * |
| Null deviance: 658.46 on 535 degrees of freedom  Residual deviance: 573.36 on 531 degrees of freedom  (31 observations deleted due to missingness)  AIC: 3006 | | | | | |

*** *p* < 0.001, ** *p* < 0.01, * *p* < 0.05

Supplemental Table 5. Summary of the optimal model (negative binomial GLM) for *CERAD Figures Copy*, predicted by the significant effect of bilingual status while accounting for education, sex, and age

| Model: glm.nb(formula = CERADFiguresCopy ~ BilingualStatus + Education + Sex + Age, link = log) | | | | | |
| --- | --- | --- | --- | --- | --- |
| Coefficients: |  |  |  |  |  |
|  | Estimate | Incidence Rate Ratio | Standard Error | z value | Pr (>\|z\|) |
| Intercept | 2.29075 | 9.88 | 0.27357 | 8.37 | < 0.001 *** |
| BilingualStatus: Active bilingual | 0.0914 | 1.10 | 0.03178 | 2.88 | **0.004** ** |
| Education | 0.0159 | 1.02 | 0.00338 | 4.7 | < 0.001 *** |
| Sex: Female | -0.04141 | 0.959 | 0.03246 | -1.28 | 0.202 |
| Age | -0.00406 | 0.996 | 0.00346 | -1.17 | 0.241 |
| Null deviance: 261.61 on 462 degrees of freedom  Residual deviance: 218.53 on 458 degrees of freedom  (104 observations deleted due to missingness)  AIC: 2079 | | | | | |

*** *p* < 0.001, ** *p* < 0.01, * *p* < 0.05

Supplemental Table 6. Summary of the optimal model (Poisson GLM) for *VOSP Number Location*, predicted by the significant interaction between bilingual status and sex while accounting for education and age

| Model: glm(formula = VOSPNumberLocation ~ BilingualStatus * Sex + Education + Age, family = “poisson”) | | | | | |
| --- | --- | --- | --- | --- | --- |
| Coefficients: |  |  |  |  |  |
|  | Estimate | Incidence Rate Ratio | Standard Error | z value | Pr (>\|z\|) |
| Intercept | 1.96172 | 7.112 | 0.29951 | 6.55 | < 0.001 *** |
| BilingualStatus: Active bilingual | -0.01801 | 0.982 | 0.05111 | -0.35 | 0.725 |
| Sex: Female | -0.26068 | 0.771 | 0.0498 | -5.24 | < 0.001 *** |
| Education | 0.02204 | 1.022 | 0.0036 | 6.12 | < 0.001 *** |
| Age | -0.00249 | 0.998 | 0.00378 | -0.66 | 0.51 |
| BilingualStatus: Active bilingual × Sex: Female | 0.14619 | 1.157 | 0.0682 | 2.14 | **0.032** * |
| Null deviance: 764.47 on 525 degrees of freedom  Residual deviance: 669.58 on 520 degrees of freedom  (41 observations deleted due to missingness)  AIC: 2569 | | | | | |

*** *p* < 0.001, ** *p* < 0.01, * *p* < 0.05

Supplemental Table 7. Summary of non-significant effects of bilingual status in neuropsychological functioning analyses

| Neuropsychological variable | Bilingual status effects: *p*-value from the likelihood ratio test of model comparisons |
| --- | --- |
| MMSE | 0.187 (χ^2^(1, *N* = 487) = 1.74, Cohen’s w = 0.06) |
| FCSRT immediate free recall | 0.594 (χ^2^(1, *N* = 529) = 0.29, Cohen’s w = 0.02) |
| FCSRT immediate cued recall | 0.702 (χ^2^(1, *N* = 529) = 0.15, Cohen’s w = 0.02) |
| FCSRT delayed free recall | 0.497 (χ^2^(1, *N* = 529) = 0.46, Cohen’s w = 0.03) |
| FCSRT delayed cued recall | 0.416 (χ^2^(1, *N* = 527) = 0.66, Cohen’s w = 0.04) |
| CERAD figures free recall | 0.456 (χ^2^(1, *N* = 464) = 0.56, Cohen’s w = 0.03) |
| Digit span backwards | 0.196 (χ^2^(1, *N* = 534) = 1.67, Cohen’s w = 0.06) |
| Semantic fluency | 0.246 (χ^2^(1, *N* = 536) = 1.35, Cohen’s w = 0.05) |
| Poppelreuter overlapping figures | 0.212 (χ^2^(1, *N* = 532) = 1.56, Cohen’s w = 0.05) |
| Neuropsychiatric inventory | 0.069 (χ^2^(1, *N* = 400) = 3.3, Cohen’s w = 0.09) |
| Geriatric depression scale | 0.954 (χ^2^(1, *N* = 528) = 0.003, Cohen’s w = 0.003) |

*Note*. *P*-values were computed from likelihood ratio tests, which were conducted to compare models with bilingual status and those without bilingual status. χ^2^ indicates the difference in deviance or log-likelihood between compared models, *N* represents the number of observations, and Cohen’s w denotes the effect size associated with the likelihood ratio test.

Supplemental Table 8. Summary of the optimal model (gamma GLM) for *CSF Aβ-42/40 ratio*, predicted by the significant interaction between bilingual status and sex while accounting for education, age, APOE status, and MMSE

| Model: glm(formula = CSFAβratio ~ BilingualStatus * Sex + Education + Age + APOE + MMSE, family = Gamma(link = “log”)) | | | | | |
| --- | --- | --- | --- | --- | --- |
| Coefficients: |  |  |  |  |  |
|  | Estimate | Rate Ratio | Standard Error | t value | Pr (>\|t\|) |
| Intercept | -3.123855 | 0.044 | 0.186157 | -16.78 | < 0.001 *** |
| BilingualStatus: Active bilingual | 0.066434 | 1.069 | 0.028905 | 2.3 | 0.022 * |
| Sex: Female | 0.037486 | 1.038 | 0.026514 | 1.41 | 0.158 |
| Education | -0.000454 | 1 | 0.001998 | -0.23 | 0.82 |
| Age | 0.000181 | 1 | 0.002077 | 0.09 | 0.93 |
| APOE44 | -0.143918 | 0.866 | 0.033545 | -4.29 | < 0.001 *** |
| APOE34 | -0.002821 | 0.997 | 0.020059 | -0.14 | 0.888 |
| APOE24 | -0.051880 | 0.949 | 0.068984 | -0.75 | 0.452 |
| APOE23 | -0.195207 | 0.823 | 0.058984 | -3.31 | 0.001 ** |
| MMSE | -0.000377 | 1 | 0.002823 | -0.13 | 0.894 |
| BilingualStatus: Active bilingual × Sex: Female | -0.093995 | 0.91 | 0.036994 | -2.54 | **0.011** * |
| Null deviance: 17.757 on 445 degrees of freedom  Residual deviance: 16.402 on 435 degrees of freedom  (41 observations deleted due to missingness)  AIC: -2977 | | | | | |

*** *p* < 0.001, ** *p* < 0.01, * *p* < 0.05

Supplemental Table 9. Summary of the optimal model (gamma GLM) for *CSF YKL-40*, predicted by the significant interaction between bilingual status and sex while accounting for education, age, and MMSE

| Model: glm(formula = CSFYKL40 ~ BilingualStatus * Sex + Education + Age + MMSE, family = Gamma(link = “log”)) | | | | | |
| --- | --- | --- | --- | --- | --- |
| Coefficients: |  |  |  |  |  |
|  | Estimate | Rate Ratio | Standard Error | t value | Pr (>\|t\|) |
| Intercept | 4.70489 | 110.486 | 0.33871 | 13.89 | < 0.001 *** |
| BilingualStatus: Active bilingual | 0.0388 | 1.04 | 0.0564 | 0.69 | 0.493 |
| Sex: Female | 0.08382 | 1.087 | 0.05363 | 1.56 | 0.121 |
| Education | 0.00201 | 1.002 | 0.00457 | 0.44 | 0.661 |
| Age | 0.01206 | 1.012 | 0.00381 | 3.16 | 0.002 ** |
| MMSE | 0.00232 | 1.002 | 0.00688 | 0.34 | 0.737 |
| BilingualStatus: Active bilingual × Sex: Female | -0.18124 | 0.834 | 0.07739 | -2.34 | **0.021** * |
| Null deviance: 6.6184 on 129 degrees of freedom  Residual deviance: 5.7985 on 123 degrees of freedom  (356 observations deleted due to missingness)  AIC: 1459 | | | | | |

*** *p* < 0.001, ** *p* < 0.01, * *p* < 0.05

Supplemental Table 10. Summary of the optimal model (gamma GLM) for *CSF GFAP*, predicted by the significant interaction between bilingual status and education while accounting for sex, age, and MMSE

| Model: glm(formula = CSFGFAP ~ BilingualStatus * Education + Sex + Age + MMSE, family = Gamma(link = “log”)) | | | | | |
| --- | --- | --- | --- | --- | --- |
| Coefficients: |  |  |  |  |  |
|  | Estimate | Rate Ratio | Standard Error | t value | Pr (>\|t\|) |
| Intercept | 5.287 | 197.744 | 1.6425 | 3.22 | 0.002 ** |
| BilingualStatus: Active bilingual | 0.9824 | 2.671 | 0.5058 | 1.94 | 0.058 |
| Education | -0.0221 | 0.978 | 0.0253 | -0.87 | 0.386 |
| Sex: Female | 0.0724 | 1.075 | 0.1889 | 0.38 | 0.703 |
| Age | 0.0325 | 1.033 | 0.0182 | 1.78 | 0.08 |
| MMSE | 0.0683 | 1.071 | 0.0308 | 2.21 | 0.031 * |
| BilingualStatus: Active bilingual × Education | -0.1141 | 0.892 | 0.0442 | -2.58 | **0.013** * |
| Null deviance: 32.762 on 58 degrees of freedom  Residual deviance: 24.938 on 52 degrees of freedom  (428 observations deleted due to missingness)  AIC: 1194 | | | | | |

*** *p* < 0.001, ** *p* < 0.01, * *p* < 0.05

Supplemental Table 11. Summary of the optimal model (gamma GLM) for *Plasma GFAP*, predicted by the significant effect of bilingual status while accounting for education, sex, age, and MMSE

| Model: glm(formula = PlasmaGFAP ~ BilingualStatus + Education + Sex + Age + MMSE, family = Gamma(link = “log”)) | | | | | |
| --- | --- | --- | --- | --- | --- |
| Coefficients: |  |  |  |  |  |
|  | Estimate | Rate Ratio | Standard Error | t value | Pr (>\|t\|) |
| Intercept | 4.7289 | 113.172 | 0.92055 | 5.14 | < 0.001 *** |
| BilingualStatus: Active bilingual | 0.21989 | 1.246 | 0.09845 | 2.23 | **0.029** * |
| Education | -0.00196 | 0.998 | 0.01116 | -0.18 | 0.861 |
| Sex: Female | 0.14521 | 1.156 | 0.10009 | 1.45 | 0.152 |
| Age | 0.01564 | 1.016 | 0.0101 | 1.55 | 0.126 |
| MMSE | -0.02139 | 0.979 | 0.017 | -1.26 | 0.213 |
| Null deviance: 12.762 on 68 degrees of freedom  Residual deviance: 10.786 on 63 degrees of freedom  (418 observations deleted due to missingness)  AIC: 833.6 | | | | | |

*** *p* < 0.001, ** *p* < 0.01, * *p* < 0.05

Supplemental Table 12. Summary of the optimal model (gamma GLM) for *CSF Aβ-42*, predicted by the significant interaction between bilingual status and sex while accounting for education, age, APOE status, and MMSE

| Model: glm(formula = CSFAβ42 ~ BilingualStatus * Sex + Education + Age + APOE + MMSE, family = Gamma(link = “log”)) | | | | | |
| --- | --- | --- | --- | --- | --- |
| Coefficients: |  |  |  |  |  |
|  | Estimate | Rate Ratio | Standard Error | t value | Pr (>\|t\|) |
| Intercept | 6.14229 | 465.118 | 0.27192 | 22.59 | < 0.001 *** |
| BilingualStatus: Active bilingual | 0.04338 | 1.044 | 0.04222 | 1.03 | 0.305 |
| Sex: Female | 0.10177 | 1.107 | 0.03873 | 2.63 | 0.009 ** |
| Education | -0.00432 | 0.996 | 0.00292 | -1.48 | 0.14 |
| Age | 0.0018 | 1.002 | 0.00303 | 0.59 | 0.554 |
| APOE44 | -0.32784 | 0.72 | 0.049 | -6.69 | < 0.001 *** |
| APOE34 | -0.05258 | 0.949 | 0.0293 | -1.79 | 0.073 |
| APOE24 | -0.3853 | 0.68 | 0.10076 | -3.82 | < 0.001 *** |
| APOE23 | -0.1185 | 0.888 | 0.08616 | -1.38 | 0.17 |
| MMSE | 0.00457 | 1.005 | 0.00412 | 1.11 | 0.269 |
| BilingualStatus: Active bilingual × Sex: Female | -0.11844 | 0.888 | 0.05404 | -2.19 | **0.029** * |
| Null deviance: 39.355 on 445 degrees of freedom  Residual deviance: 33.774 on 435 degrees of freedom  (41 observations deleted due to missingness)  AIC: 5759 | | | | | |

*** *p* < 0.001, ** *p* < 0.01, * *p* < 0.05

Supplemental Table 13. Summary of non-significant effects of bilingual status in biomarker analyses

| Biomarker variable | Bilingual status effects: *p*-value from the likelihood ratio test of model comparisons |
| --- | --- |
| CSF Aβ-40 | 0.131 (χ^2^(1, *N* = 446) = 0.18, Cohen’s w = 0.02) |
| CSF p-tau 181 | 0.35 (χ^2^(1, *N* = 449) = 0.22, Cohen’s w = 0.02) |
| Plasma p-tau 181 | 0.558 (χ^2^(1, *N* = 84) = 0.07, Cohen’s w = 0.03) |
| Plasma p-tau 217 | 0.112 (χ^2^(1, *N* = 69) = 0.5, Cohen’s w = 0.09) |
| CSF t-tau | 0.458 (χ^2^(1, *N* = 448) = 0.11, Cohen’s w = 0.02) |
| CSF NfL | 0.934 (χ^2^(1, *N* = 127) = 0.002, Cohen’s w = 0.004) |
| Plasma NfL | 0.253 (χ^2^(1, *N* = 169) = 0.32, Cohen’s w = 0.04) |

*Note*. *P*-values were computed from likelihood ratio tests, which were conducted to compare models with bilingual status and those without bilingual status. χ^2^ indicates the difference in deviance between compared models, *N* represents the number of observations, and Cohen’s w denotes the effect size associated with the likelihood ratio test.
